# Replacement of the Gamma by the Delta variant in Brazil: impact of lineage displacement on the ongoing pandemic

**DOI:** 10.1101/2021.12.27.21268309

**Authors:** Marta Giovanetti, Vagner Fonseca, Eduan Wilkinson, Houriiyah Tegally, Emmanuel James San, Christian L. Althaus, Joilson Xavier, Svetoslav Nanev Slavov, Vincent Louis Viala, Alex Ranieri Jerônimo Lima, Gabriela Ribeiro, Jayme A. Souza-Neto, Heidge Fukumasu, Luiz Lehmann Coutinho, Rivaldo Venancio da Cunha, Carla Freitas, Carlos F Campelo de A e Melo, Wildo Navegantes, Rodrigo Fabiano do Carmo Said, Maria Almiron, Tulio de Oliveira, Sandra Coccuzzo Sampaio, Maria Carolina Elias, Dimas Tadeu Covas, Edward C. Holmes, Jose Lourenço, Simone Kashima, Luiz Carlos Junior de Alcantara

## Abstract

The COVID-19 epidemic in Brazil was driven mainly by the spread of Gamma (P.1), a locally emerged Variant of Concern (VOC) that was first detected in early January 2021. This variant was estimated to be responsible for more than 96% of cases reported between January and June 2021, being associated with increased transmissibility and disease severity, a reduction in neutralization antibodies and effectiveness of treatments or vaccines, as well as diagnostic detection failure. Here we show that, following several importations predominantly from the USA, the Delta variant rapidly replaced Gamma after July 2021. However, in contrast to what was seen in other countries, the rapid spread of Delta did not lead to a large increase in the number of cases and deaths reported in Brazil. We suggest that this was likely due to the relatively successful early vaccination campaign coupled with natural immunity acquired following prior infection with Gamma. Our data reinforces reports of the increased transmissibility of the Delta variant and, considering the increasing concern due to the recently identified Omicron variant, argues for the necessity to strengthen genomic monitoring on a national level to quickly detect and curb the emergence and spread of other VOCs that might threaten global health.

## Main text

Since late 2020 the evolution of SARS-CoV-2 has been characterised by the appearance of mutations on the Spike protein leading to the emergence of so-called ‘variants of concern’ (VOC) some of which have spread globally^1,2^. In particular, some of the mutations in VOCs are suggested to impact viral transmissibility^3^, resistance to neutralizing antibodies^4^, and virulence. The identification of such variants has recently challenged public health authorities with respect to tracking transmission and mitigating the impact in the ongoing pandemic. To date, the most important VOCs documented are Alpha, Beta, Gamma, Delta and Omicron (described in late November 2021), first detected in United Kingdom, South Africa, Brazil, India, and South Africa/Botswana respectively^5–8^.

In the context of continuous surveillance of SARS-CoV-2 clinical samples on behalf of the National Pandemic Alert Network based in the State of São Paulo (Brazil) since early January 2021, our team has been monitoring the proportion of circulating variants in Brazil. Data from this initiative shows that after July 2021 the Delta variant has become highly prevalent. It has been suggested that the Delta variant might be more transmissible^9^, having led to public health emergencies in other countries due to overwhelming increases in the number of cases, hospitalizations and deaths when compared to previous circulating variants. Here we describe how Delta became the predominant SARS-CoV-2 variant in Brazil, rapidly replacing the previously dominant Gamma, and how this displacement was not associated with an increase in reported case numbers or deaths.

The COVID-19 epidemic in Brazil can generally be characterized by two epidemic waves accounting for more than 22 million cases and 616,251 deaths until early December 2021^10^. The first epidemic wave was characterised by the circulation of multiple SARS-CoV-2 lineages (among them the B.1.1.28 and the B.1.1.33), as a direct consequence of multiple independent introduction events between February and March 2020 (Fig.1A)^11^. By the end of October 2020, even with the implementation of non-pharmaceutical interventions (NPIs), a second wave associated with a dramatic resurgence in cases and death numbers took place. This wave was fuelled by the emergence and circulation of several Variants under Monitoring (VUM), such as P.2 (i.e., Zeta), and some VOCs including Alpha and later Gamma (i.e., P.1), which became widespread by January 2021 and dominated the viral population for nearly 8 consecutive months (Figure 1A). Despite the national vaccination rollout beginning on January 17th 2021, the COVID-19 death toll in the country steadily rose in March 2021, reaching a peak in April 2021 (Fig. 1A). This was followed by a decrease in the number of daily cases and deaths by April (Fig. 1A and Fig. S1), likely as a consequence of a gradual increase in population immunity. Zeta (P.2) mostly dominated the first epidemic wave persisting up to March 2021 when it was replaced by Gamma (P.1) (Fig. 1C). This period was characterized by an upsurge in the number of total cases with a peak registered between February and June 2021. The Alpha variant was also detected from January 2021 onwards, but it remained at a very low frequency nationally (less than 2%)^12^.

**Figure 1.**
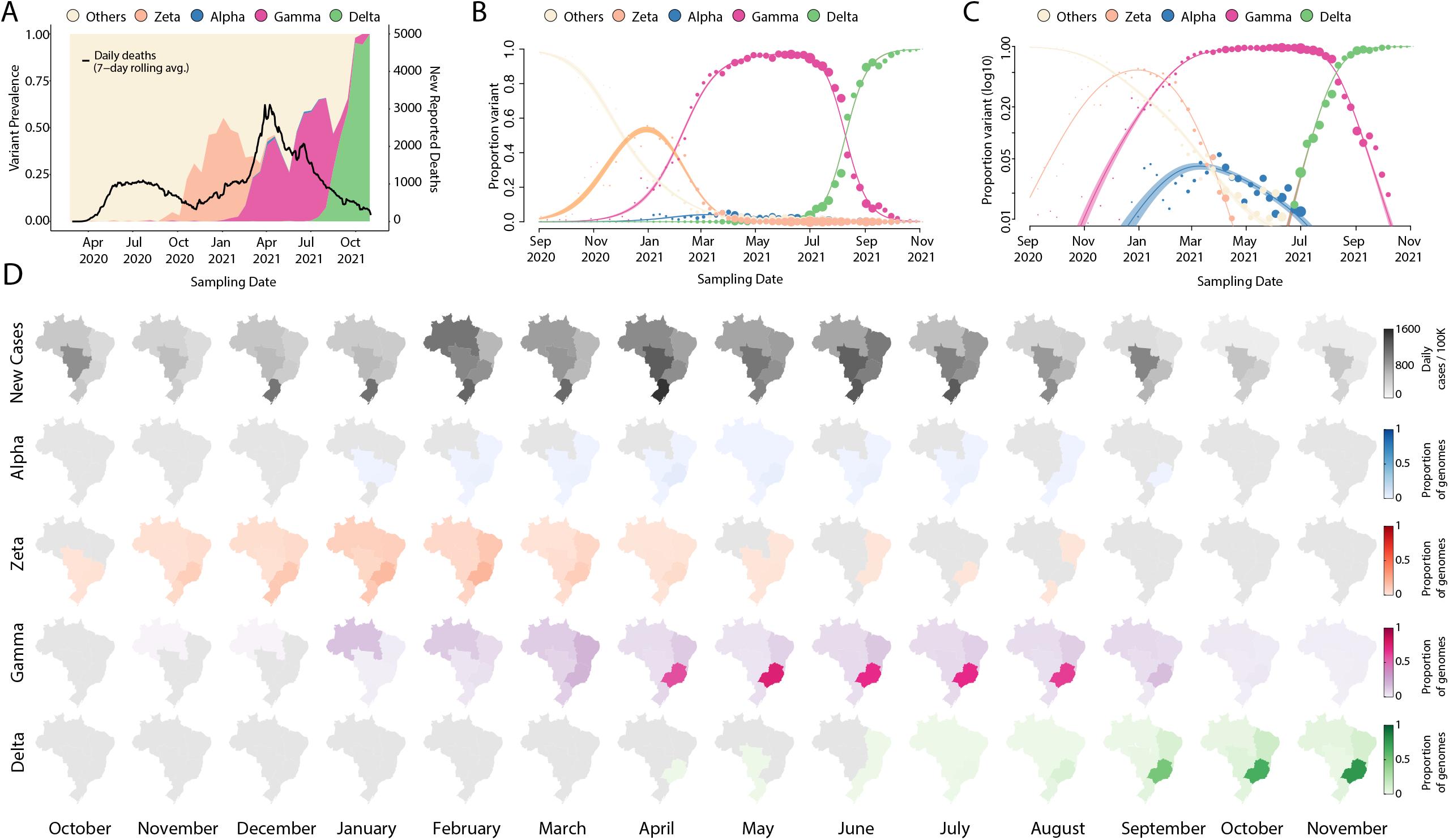
Replacement of Gamma by Delta variant in Brazil. (A) Dynamics of the SARS-CoV-2 epidemic in Brazil showing the number of daily COVID-19 deaths and the progression in the proportion of circulating variants in the country over time, with the rapid replacement of the Gamma by the Delta variant. (B-C) Modelled proportion of SARS-CoV-2 variants over time in Brazil in a linear and in a logarithmic scales, showing that Gamma became the dominant variant in Brazil by the beginning of 2021 and was rapidly outcompeted by Delta from June 2021. Model fits are based on a multinomial logistic regression. The size of dots corresponds to the weekly sample size. (D) Prevalence maps following the progression in the monthly average of daily number of cases and proportions of relevant variants per region (North, Northeast, Midwest, Southeast and South) in Brazil from October 2020 to October 2021.

In April 2021, through intensified sampling of likely imported cases associated with returning travellers, the Delta variant was detected. Delta first spread in the southeast, the most populous region of Brazil harbouring the largest urban centres and airports with highest national and international travel flows, that continued to operate throughout 2021^13^. Throughout July 2021, Delta rapidly displaced Gamma, becoming the dominant variant circulating nationally during August 2021 (Fig. 1A and Fig. S1). This variant displacement was not associated with a concurrent increase in COVID-19 incidence (Fig. 1C). We estimate that Delta had a growth advantage of 0.064 (95% confidence interval [CI] 0.058-0.071) per day compared to Gamma (Fig. 1B). Assuming that the variants have the same generation time of 5.2 days^14^, this corresponds to a growth advantage of 33% (95% CI 30-37%) per generation of viral transmission, which is in good agreement with earlier findings based on sequence data from multiple countries^15^.

Although the precise cause of these trends is unknown, we hypothesize that a relatively successful early vaccination campaign in the region coupled with a reasonable percentage of the population with natural immunity acquired by prior infection with Gamma contributed to the decreasing case rate around the time of introduction of the Delta variant. In contrast, when Gamma dominated transmission, the observed increasing in the number of cases would have been attributed to a relatively naïve population to infection with any SARS-CoV-2 variant. This epidemiological scenario of rapid switching of new variants without an increase in the number of cases seems not to be uncommon and has also been observed elsewhere^16–20^.

We further estimated phylogenetic trees to explore the relationship of the sequenced Brazilian genome to those of other isolates across the world. For this purpose, we retrieved 11,147 Delta genomes from Brazil, from which 6,626 were generated by our National Pandemic Alert Network and a globally representative set of other Delta genomes (n=13,261). Our time-stamped phylogeny revealed that the Brazilian Delta isolates are scattered throughout the phylogeny suggesting multiple independent introductions (Fig.2A, Figure S2).

Our analysis revealed at least 124 independent introductions of the Delta variant into Brazil between October 2020 and October 2021 (Fig 2b), with the majority (30%) originating from North America, followed by India (17%), European countries (mostly UK 13%), and other South American countries.

**Figure 2.**
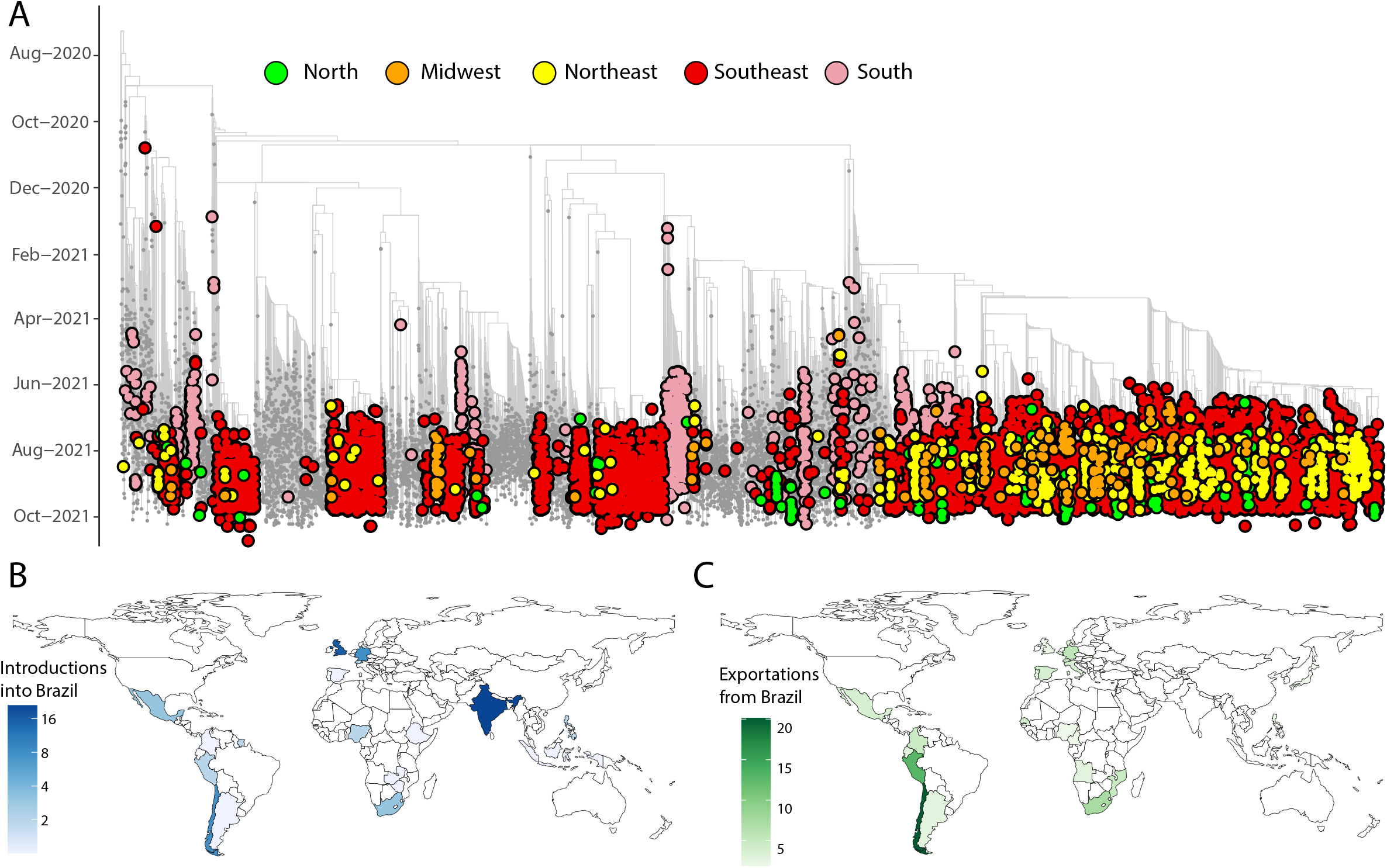
Phylogenetic analysis of the Delta variant in Brazil. (A) Time-resolved maximum clade credibility phylogeny of 11,147 SARS-CoV-2 Delta sequences from Brazil, from which 6,626 were generated by our National Network for Pandemic Alert of SARS-CoV-2 along with global Delta sequence (grey). Colours indicate different Brazilian regions (North in green; Midwest in orange; Northeast in yellow; Southeast in red and South in light pink); (B) Inferred locations of importations of the Delta variant into Brazil. (C) Inferred locations of exportation of the Delta variant from Brazil.

Following introductions into Brazil, in line with recent findings^21^, the Delta variant appears to have further spread within Brazilian regions (Fig.2A), highlighting complex local transmission dynamics maintained by travel. Importantly, this variant appears to have been introduced into each region of Brazil through multiple ports of entry making it challenging to accurately reconstruct transmission pathways across the country. Our analysis further shows that the Delta variant subsequently spread from Brazil into countries in South America, and also to North America, Europe and Africa (Fig 2C)^22^.

The first months of the Brazilian epidemic were fuelled by the circulation of multiple lineages, as a direct consequence of multiple viral independent introductions from overseas. As the epidemic progressed, the observed large-scale community transmission led to local emergence of VOCs and VUMs^23^. These variants dominated the end of the first and second epidemic wave in the country, resulting in an exponential increase in the number of daily cases and deaths, making Brazil one of the countries hardest hit by the severe acute respiratory syndrome coronavirus 2 (SARS-CoV-2) pandemic worldwide.

In this study we analysed the displacement of Gamma by the Delta variant in Brazil, showing also how this replacement was not associated with an increase in reported cases numbers. Given that during the displacement NPIs remained relatively relaxed locally, it seems likely that a significant proportion of the Brazilian population has developed immunity (either through natural infection or vaccination) which helped to prevent a rise in case numbers. We estimated that in Brazil, Delta had a transmission advantage of 33% (95% CI 30-37%) compared to Gamma. The spread of future variants will be possible if they exhibit increased transmissibility or immunity evasiveness, both of which seem likely in the case of the Omicron variant^8^. However, as shown here, displacement does not necessarily equate to new epidemic waves of reported cases or deaths, since the outcome of the spread of a new variant will depend on a complex interplay between local interventions, infrastructure and the immunity landscape already present in the population due to the circulation of previous variants. Considering that past replacement events have taken place so rapidly in Brazil, future epidemiological assessment of new VOC must be conducted rapidly and regularly. The capacity to detect and respond to new variants requires continued support and funding for molecular surveillance and sequencing capacity more generally. Moreover, enhanced sampling efforts in countries like Brazil are needed to ensure better geographical representativeness of available SARS-CoV-2 sequences and the rapid detection of emerging variants when their frequencies are still low. These factors will be key to detect, understand and respond to the likely upcoming Omicron wave which has already been detected in more than 87 countries, including Brazil, and which will possibly alter the landscape of variants currently circulating in the country.

## Data Availability

All SARS-CoV-2 whole genome sequences produced by the National Network for Pandemic Alert of SARS-CoV-2 have been deposited in the GISAID sequence database and are publicly available subject to the terms and conditions of the GISAID database. The GISAID accession numbers of sequences used in the phylogenetic analysis, are provided in the Supplementary Table S1. https://www.gisaid.org/

## Data availability

All SARS-CoV-2 whole genome sequences produced by the National Network for Pandemic Alert of SARS-CoV-2 have been deposited in the GISAID sequence database and are publicly available sub- ject to the terms and conditions of the GISAID database. The GISAID accession numbers of sequenc- es used in the phylogenetic analysis, are provided in the **Supplementary Table S1**.

## Code availability

All input files along with all resulting output files and scripts used in the present study will be made available upon request and publicly shared on GitHub at final publication.

## Acknowledgments

The authors acknowledge the National Network for Pandemic Alert of SARS-CoV-2 and the contribution of all employees of General Coordination of Public Health Laboratories and professionals of Public Health Laboratories of Brazil for their contribution towards the sequencing effort and for their commitment and work during the fight of the COVID-19 pandemic. We also thank all the authors who have kindly deposited and shared genome data on GISAID.

## Author Contributions

Molecular screening and produced SARS-CoV-2 genomic data: MG, JX, SNS, VLV, ARJ, GR, JASN, HF, LLC, SCS, MCE, and SK; Collected samples and curated metadata: SCS, MCE, VF, DTC, SK, LCJA, CF, CFCAM, WN, RFCS, MA; Analysed the data: MG, VF, EW, HT, EJS, CLA; Helped with study design and data interpretation: MG, VF, EW, HT, EJS, CLA;, ECH, JL, and LCJA; Wrote the initial manuscript, which was reviewed by all authors: MG, CLA, EH, JL and LCJA.

## Materials and Methods

### Ethics statement

This research was approved by the Ethics Review Committee of the Pan American Health Organization (PAHOERC.0344.01) and by the Federal University of Minas Gerais (CEP/CAAE: 32912820.6.1001.5149). The availability of these samples for research purposes during outbreaks of national concern is allowed under the terms of the 510/2016 Resolution of the National Ethical Committee for Research – Brazilian Ministry of Health (CONEP - Comissão Nacional de Ética em Pesquisa, Ministério da Saúde) that authorize, without the necessity of an informed consent, the use of clinical samples collected in the Brazilian Central Public Health Laboratories to accelerate knowledge building and contribute to surveillance and outbreak response. The samples processed in this study were obtained anonymously from material exceeding the routine diagnosis in Brazilian public health laboratories that belong to the public network within BrMoH.

### Epidemiological data

We analysed daily cases of SARS-CoV-2 in Brazil up to 2^nd^ November 2021 from the COVIDA network available at https://github.com/wcota/covid19br. For convenience, the geographical locations were aggregated by Brazilian macro regions: North, Northeast, Southeast, South, and Midwest. The National COVIDA network releases daily updates on the number of confirmed new cases, deaths, and recoveries, with a breakdown by states and regions.

### Sample collection and molecular diagnostic assays

As part of the National Pandemic Alert Network, since early January 2021, our team has been monitoring the proportion of circulating variants in Brazil. For this purpose, convenience clinical samples, from public laboratories in Brazil were received and randomly selected for sequencing every week. Depending on the partner institution, library preparation and sequencing was done either on the Illumina and/or Oxford Nanopore Platform. Viral RNA was extracted from nasopharyngeal swabs using an automated protocol and tested for SARS-CoV-2 by multiplex real-time PCR assays: (i) the Allplex 2019-nCoV Assay (Seegene) targeting the envelope (E), the RNA dependent RNA polymerase (RdRp) and the nucleocapsid (N) genes; (ii) the Charité: SARS-CoV2 (E/RP) assay (Bio-Manguinhos/Fiocruz) targeting the E gene, and (iii) the GeneFinder COVID-19 Plus RealAmp Kit (Osang Healthcare, South Korea) supplied by the Brazilian Ministry of Health (BrMoH), Butantan Institute and the Pan-American Health Organization (OPAS).

### cDNA synthesis and whole genome sequencing

Samples were selected for sequencing based on the Ct value (≤30) and availability of epidemiological metadata, such as date of sample collection, sex, age and municipality of residence. The preparation of SARS-CoV-2 genomic libraries was performed using two different strategies: (i) the Illumina COVIDSeq test following the manufacturer’s instructions^11^; and (ii) the Oxford Nanopore sequencing using the ARTIC Network primal scheme^24^. The normalized libraries were loaded for the Illumina sequencing onto a 300-cycle MiSeq Reagent Kit v2 and run on the Illumina MiSeq instrument (Illumina, San Diego, CA, USA) and for the Nanopore strategy into a R9.4 flow cell (Oxford Nanopore Technologies) as previously described^25^. All experiments were performed in a biosafety level-2 cabinet. In each sequencing run, we used negative controls to prevent and check for possible contamination with less than 2% mean coverage.

### Genome assembly

Sequences generated on the Illumina and nanopore platforms were assembled using Genome Detective 1.132/3^26^.

### Estimating relative transmission advantage

We analyzed 11,147 SARS-CoV-2 Brazilian sequences, from which n=6,626 were generated by our National Pandemic Alert Network, that have been uploaded to GISAID (Global Initiative On Sharing All Influenza Data) from 26 April 2021 to 23 October 2021. We used a multinomial logistic regression model to estimate the growth advantage of Delta compared to Gamma in Brazil^15,27^ We added splines to account for time-varying growth rates in the model fit and estimated the overall growth advantage without splines. We fitted the model using the *multinom* function of the *nnet* package (Venables & Ripley)^28^ in R.

### Phylogenetic analysis

We analyzed 11,147 Delta variants from Brazil, publicly available on GISAID^21^ as at the 4 November 2021, of which n=6,626 were generated by our National Pandemic Alert Network and used in this study. These were put in their phylogenetic context through comparison with a globally representative (n=13,261) set of other SARS-CoV-2 Delta variants from around the world, sampled from October 23th 2020 to October 23th 2021. The full set of sequences were aligned with NextAlign^29^ to obtain a good codon quality alignment against the Wuhan-Hu-1 universal reference sequence. The subsequent alignment was then used to infer a Maximum Likelihood tree topology in IQTREE2^30^ (-m GTR, -b 100). Transfer bootstrap support for splits in the topology was inferred using Booster^31^. The resulting consensus ML tree topology was assessed for molecular clock signal in TempEst^32^. Potential outlier sequences or sequences lacking required metadata (e.g. date and location of sampling) were pruned off the topology with the ape package^33^ in R prior to dating. The branches in the ML-tree topology were then converted into units of calendar time in TreeTime^34^ using a constant rate of 0.0008 substitutions/site/year^35^ with a clock standard deviation of 0.0004 substitutions/site/year. Following the dating of the phylogeny we annotated the tips and internal nodes using the “mugration” package extension of TreeTime and then counted the state changes from one country to another and their inferred time points. This gave us the number and time of SARS-CoV-2 Delta isolates entering Brazil over time. To obtain a measure of confidence in the time and source of viral transitions, we performed the discrete ancestral state reconstruction on 10 bootstrap replicate trees.

## Additional information

**Supplementary Figure 1**. Dynamics of the SARS-CoV-2 epidemic in each Brazilian showing the number of daily COVID-19 deaths and the progression in the proportion of circulating variants in the country over time, showing the rapid replacement of Gamma by Delta.

**Supplementary Figure 2**. Progressive distribution of the top 10 Delta PANGO sub-lineages in Brazil over time.

**Supplementary Table 1**. GISAID accession numbers of sequences used in the phylogenetic analysis.

