## Supplementary figures and images for "Replacement of the Gamma by the Delta variant in Brazil: impact of lineage displacement on the ongoing pandemic"

### Supplementary_Figure_1

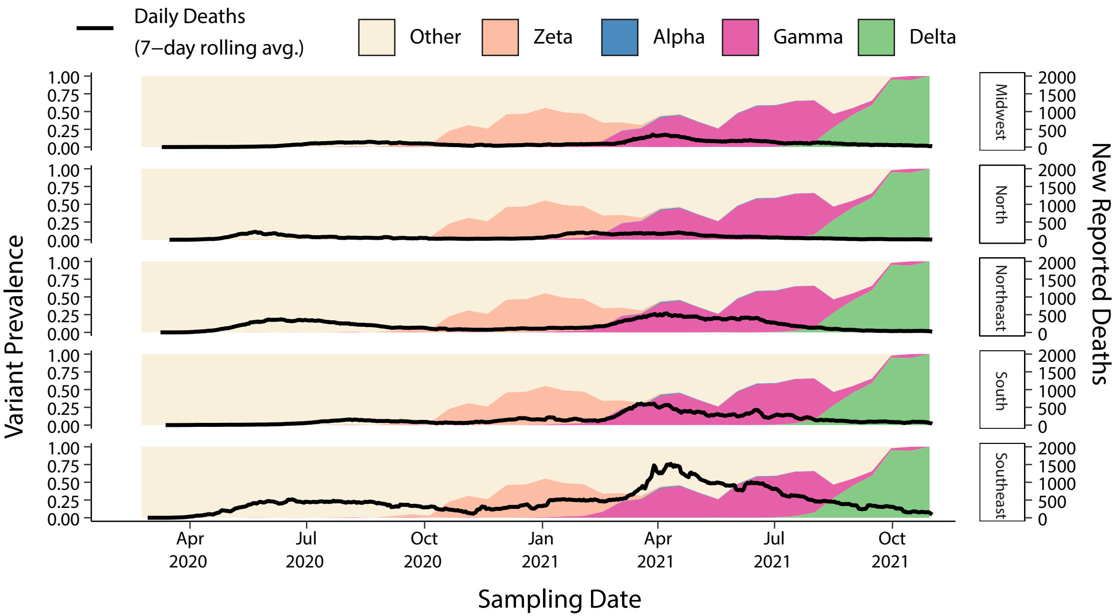

### Supplementary_Figure_2

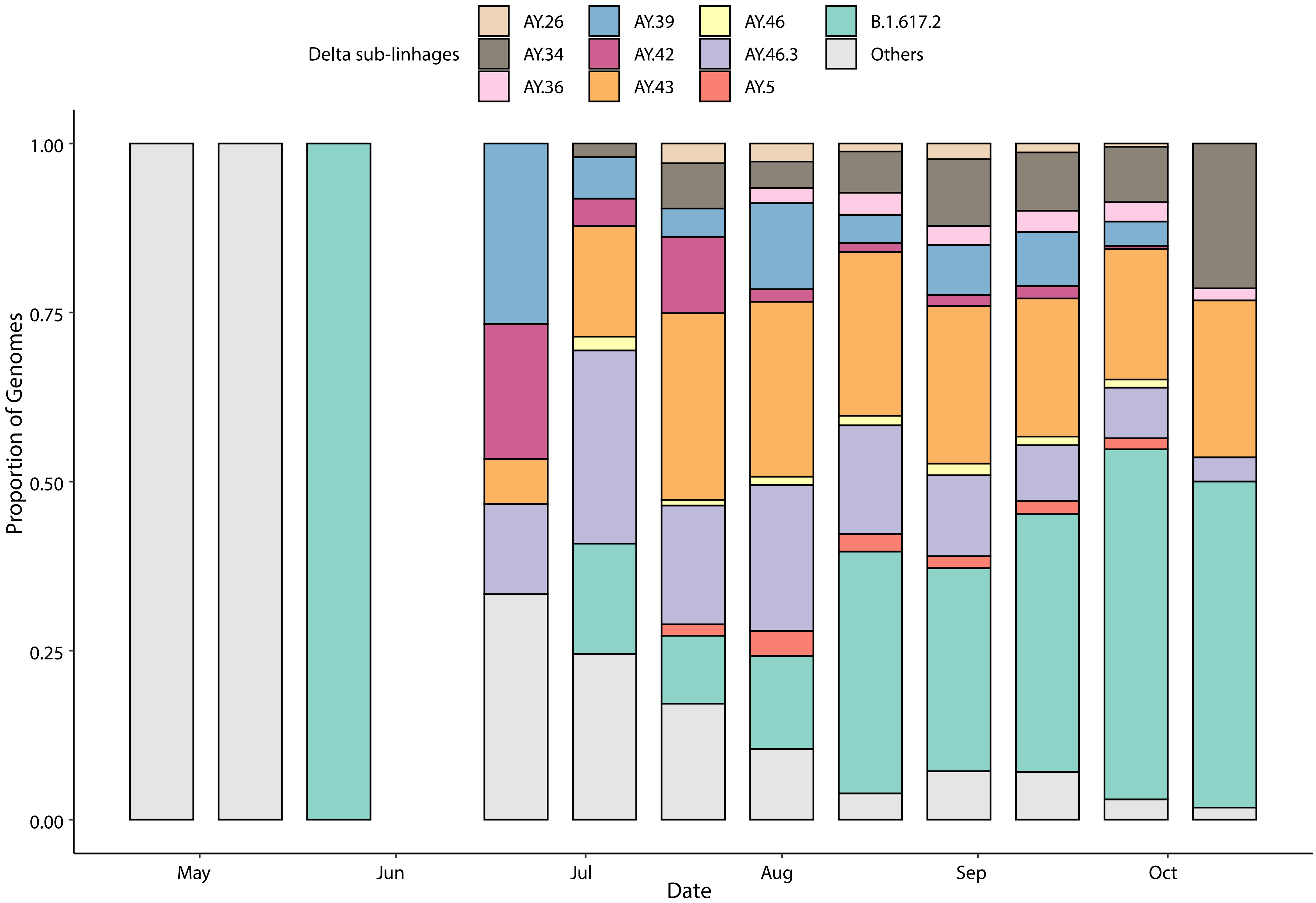
